# Co-designed point-of-care ultrasound program development and implementation at a Veterans Affairs Emergency Department

**DOI:** 10.1101/2024.12.04.24318469

**Authors:** Rebecca G. Theophanous, Catherine A. Staton, Luna C. Ragsdale, Anna Tupetz, Erica Peethumnongsin, Stephanie A. Eucker

**Affiliations:** Department of Emergency Medicine, Duke University School of Medicine, Durham, NC 27710, USA; Durham Veterans Affairs Healthcare System, Durham, NC 27710, USA; Duke Global Health Institute, Duke University, Durham, NC 27710, USA

**Keywords:** point-of-care ultrasound (POCUS), experience-based co-design, participatory action research, end-user engagement, healthcare innovation, Veterans Affairs healthcare system

## Abstract

Integrating point-of-care ultrasound (POCUS) into patient care requires a multi-faceted culture change using a multi-pronged approach. A standardized implementation method is lacking for POCUS program sustainability. We developed a standardized training curriculum and clinical POCUS documentation, archival, and image review process at a single Veterans Affairs Emergency Department (ED). We hypothesized that co-designed development and implementation of the multifaceted POCUS intervention would maximize ED provider uptake and sustainability.

Using Participatory Action Research (PAR) and adapted Experience-Based Co-design (EBCD) methods, stakeholders collaboratively co-designed our POCUS intervention to optimize implementation. Using the PRODUCES framework, twelve stakeholder participants (co-creators including study researchers, ultrasound faculty, ED leadership, and ED providers (POCUS end-users)) met in four monthly co-design meetings from July to October 2021 to brainstorm, discuss, refine, and finalize the POCUS intervention and implementation plan.

Throughout the co-design process, stakeholders reviewed findings from prior meetings, reflected on successes and failures, and held open discussions on refining and finalizing the proposed POCUS educational and clinical program. By involving stakeholders as co-creators throughout the co-design process, we maximized end-user POCUS enthusiasm, program uptake, and sustained use. This simple, streamlined, and generalizable user-centered co-design method serves as a framework for future POCUS implementation and dissemination plans.

## I. Introduction

Point-of-care ultrasound (POCUS) augments diagnostic and procedural clinical patient care in emergency departments (ED) and hospitals nationwide as a low-cost, safe, and increasingly available bedside tool.[1,2] ED providers are frequent POCUS users and are at the forefront of POCUS education and innovation, with prior ED studies showing that POCUS use expedites bedside care and improves patient satisfaction and clinical outcomes.[1,2] However, a sustainable POCUS program requires a significant multi-faceted procedural and cultural change for new users--including incorporating regular POCUS use into clinical practice, and implementation of standardized education, image documentation, storage, and quality review processes.[3,4] Gaps in POCUS training and lack of program infrastructure still exist, and a standardized user-centered implementation plan remains elusive, particularly among community EDs and within the Veterans Affairs (VA) healthcare system.[5–8] The VA healthcare system is the largest integrated healthcare system in the United States and serves over 2 million veterans in EDs and urgent cares annually. Nationwide, these settings frequently comprise heterogenous groups of providers from different medical specialties (e.g., emergency medicine, internal medicine, and family medicine), training backgrounds (e.g., MD, DO, NP, and PA), and clinical practice environments (e.g. rural, urban, different patient volumes), thus is difficult to tailor an educational or clinical intervention toward the whole group rather than specific subgroups.

A potential solution for improving healthcare processes and systems currently championed by the VA is the use of co-design for developing sustainable systems.[9–11] Co-design in healthcare research is defined as “*the meaningful involvement of research users during the study planning phase of a research project*”.[12] In healthcare research studies, Participatory Action Research (PAR) involves collaboration between researchers and end-users to understand and change systems through active end-user involvement.[12,13] PAR increases the frequency and intensity of end-user engagement and can yield positive feedback and growth among end-users.[9,12] Applied to POCUS, a systems-based cultural change, rather than a one-way POCUS intervention created by ultrasound faculty for ED providers, must be implemented to achieve an effective and sustainable solution. In addition, we used Evidence-Based Co-Design (EBCD) methods which emphasize setting clear participant expectations and assigning defined participant roles.[13–17] In this way, stakeholder involvement in project design and adaptation aims to achieve maximum acceptability and effectiveness specific to both stakeholders and setting.[9,10] Furthermore, end-user participation through EBCD changes clinical and educational practice through experience-based knowledge. As the implementation strategy components are integrated and grow over a longitudinal timeline, we can continuously evolve and improve our POCUS intervention through repetitive iterative cycles of performance and reflection.[9,13,15,18]

Therefore, our aim was to create and implement a multifaceted POCUS intervention involving a standardized POCUS educational curriculum, clinical documentation and archiving system, image review and quality improvement process, and department-wide culture change amongst ED providers at a single site VA ED using PAR and adapted EBCD methods.[9,14–17] Through co-design, our goal was to maximize POCUS uptake and sustainability for users with a wide range of baseline POCUS knowledge and skills, and to develop methods that can serve as a framework for future POCUS implementation efforts.

## II. Methods

### 2.1 Study design

We used co-design PAR and adapted EBCD methodology at the VA ED implementation site to develop and refine a multifaceted POCUS intervention for ED providers that would include both an educational training program and a clinical implementation program with documentation, archiving, image review, and quality improvement processes.[9,12–15,19] Study researchers used the PRODUCES (Problem, Objective, Design, end-Users, Co-creators, Evaluation, Scalability) framework to frame the study aim and identify problems with the POCUS program intervention and implementation strategy (Table 1).[9,14,18] We used the GRIPP2 Long Form for co-designed studies and the Checklist for Reporting Intervention Co-creation.[9,20] Our study was deemed exempt by the VA Institutional Review Board, and consent was waived by the IRB (1631300-4).

**Table 1:**
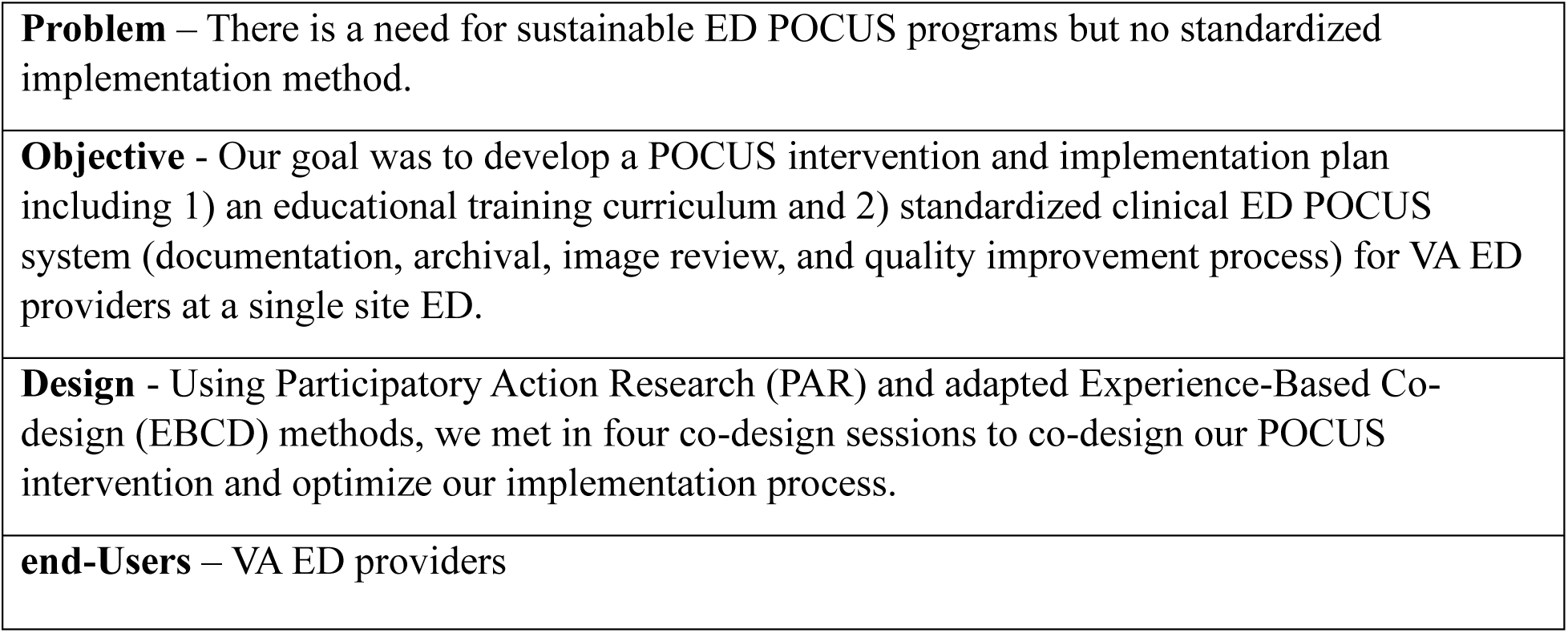
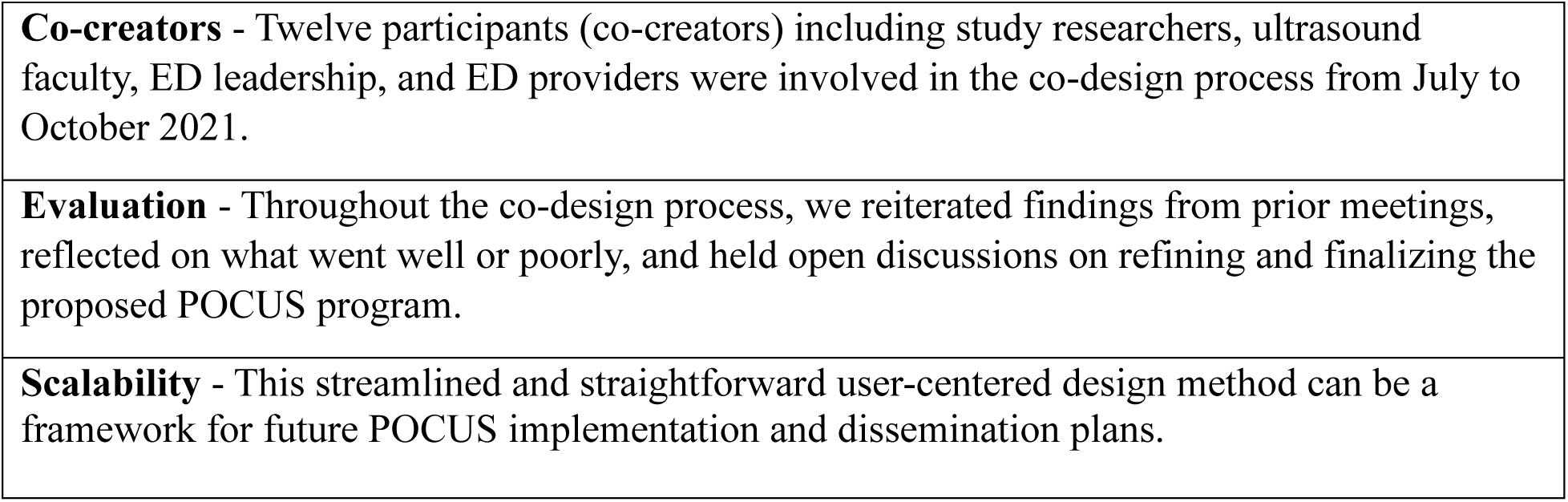
PRODUCES framework.

### 2.2 Study Setting

The primary performance site was a 251-bed tertiary care referral, teaching, and research hospital in the VA healthcare system (VAHCS) affiliated with a local academic medical center. The VAHCS ED evaluates and treats approximately 26,000 patients annually.

### 2.3 Study participants and recruitment

We invited primary ED providers, which were those ED providers who worked in the ED on a regular basis (24 attendings and 1 APP). We followed a convenience sampling strategy and recruited participants via word of mouth, emails to the full ED provider list, and announcements at monthly VA ED provider meetings. We also invited VA ED leadership and lead educational faculty who were interested in the POCUS implementation process. We excluded nurses and intermediate care technicians (ICTs or former military corpsmen or medics working in the ED) as their ultrasound training and use differs from that of primary ED providers. Table 2 describes the study inclusion and exclusion criteria. Table 3 describes the stakeholder groups included as co-creators in the co-design process. Study participation was voluntary. Co-creators were all equally involved in the co-design process to represent their viewpoints and were responsible for providing input from their respective groups to manifest ownership. These groups include the academic research team, ultrasound faculty, VA ED leadership, and primary ED providers (end-users) and together comprise the co-creator team.

**Table 2:** Inclusion and exclusion criteria for the VA POCUS intervention co-design sessions.

| Included (n=total within the group) | Excluded |
| --- | --- |
| Research team (n=11) | ED nurses |
| Ultrasound faculty (n=2) | Intermediate Care Technicians (ICTs) |
| ED leadership (ED chief, Deputy Director, committee and education leads) (n=4) | Ancillary ED providers (14 moonlighting fellows) |
| Primary ED providers (24 attendings, 1 APP) |  |

**Table 3:** List of stakeholder groups included as co-creators in the co-design process.

| Group | Definition | Example | Expertise |
| --- | --- | --- | --- |
| Academic research team (n=4) | Individuals who conduct the research study | University researchers, study mentors | Research knowledge from prior experience, literature review, and mentorship |
| Ultrasound faculty (n=1) | Individuals who are interested in the implementation intervention and are experts on ultrasound education and clinical use | Ultrasound-trained ED physicians | Knowledge experts on ultrasound literature, POCUS education, and clinical POCUS system processes |
| VA ED leadership (n=3) | Administrative ED faculty who make decisions on behalf of the group, meet with other hospital groups for collaboration and cohesive medical care, and could potentially benefit from the intervention's success | ED Chief, Deputy Director, lead educational faculty | Provide insight on the intervention development and have experience with the ED and VA hospital system processes and resources, including personnel |
| ED providers (end-users) (n=4) | Group or individuals who are the target of the intervention and could potentially benefit from its success | Primary ED providers (ED attendings and APP) | Provide insight on specific group and individual needs |

N=number of co-creator participants from each group

### 2.4 Co-design process: identifying co-design goals, topics, and relevant background information

Using PRODUCES, the <u>P</u>roblem was a need for a sustainable ED POCUS program and lack of a standardized implementation system. The goal or <u>O</u>bjective of the co-design meetings was to develop 1) a curriculum of topics to be covered in educational sessions, and 2) optimal processes for the clinical POCUS system (documentation, archiving, image review, equipment maintenance, and quality improvement and program feedback). Ultrasound faculty and the research team performed a literature review to understand what types of POCUS education or clinical interventions had been implemented at other sites, their effectiveness, and evaluation process.

### 2.5 Structure of the co-design sessions (Figure 1)

We convened in four monthly co-design meetings from July to October 2021 to analyze and provide direct feedback on further improving our POCUS intervention components (Design). Each individual’s expertise (<u>C</u>o-creators) was elicited to maximize the impact of our intervention’s development and sustainability. For example, ultrasound faculty researched and proposed educational material for the teaching curriculum through review of published ultrasound studies and validated curriculums. They then tailored topics toward knowledge and skills gaps identified by <u>E</u>nd-users, who were represented by the co-creators. Session formats, timing, group sizes, and other details were modified based on input from VA ED leadership and education leads.

**Figure 1:**
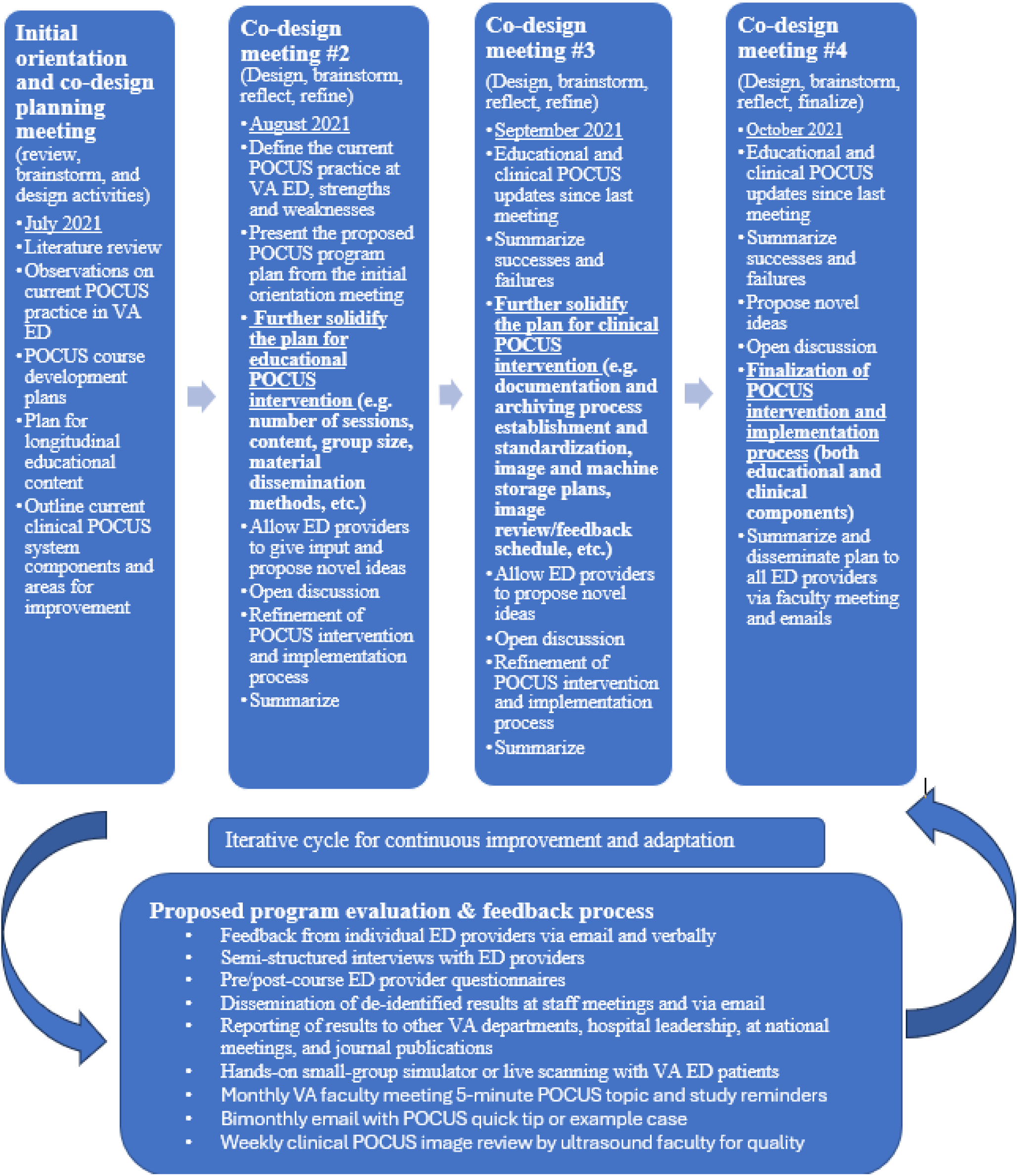
VA POCUS program Co-design Process and Timeline

#### 1. Initial orientation and co-design planning meeting #1

For our POCUS intervention and strategic implementation, we first framed the problem by holding an initial co-design orientation meeting to define and discuss key stakeholder requirements amongst the co-creator groups. We oriented co-creators to the study and the proposed POCUS implementation intervention and plan, strategized methods for deployment and information dissemination, and planned the initial designs of the educational and clinical program components.

For POCUS education, we outlined a longitudinal POCUS curriculum involving both hands-on education and online/textbook ultrasound resources. We proposed offering an introductory basic skills POCUS course and smaller monthly hands-on POCUS sessions, with ultrasound faculty and skilled POCUS users as instructors. Next, we planned what specific educational content to include and mapped out the course structure.

For the clinical POCUS systems intervention, ultrasound faculty and ED leadership worked together to lay out a framework for required components including ultrasound documentation, archiving, and image review. ED leadership assisted in identifying appropriate contacts from other departments including BioMedical and support staff to understand how to implement a program within our ED specifically (e.g. internet connections and network capabilities, setting up an ultrasound procedure order in the electronic health record (EHR), POCUS exam template notes, etc.).

#### 2. Co-design meetings #2-4

Three additional co-design meetings were held monthly via secured video conferencing and lasted approximately one hour, incorporating member checking and respondent validation. The co-design meetings were structured to focus on refining 1) the educational POCUS component, 2) the clinical POCUS component, and 3) finalization of both components, but also intentionally included topic overlap for cyclical improvement. For each meeting, we summarized prior work, recapped the project status, allowed for open discussion and new ideas in a safe, protected space, time for reflection and feedback on processes that were going well or poorly, and reiterated key points and future tasks or goals at the meeting’s conclusion (<u>E</u>valuation).

To start each meeting, the overall aim and session’s purpose were highlighted. End-users were up-skilled or given an overview of co-design methodology and required POCUS system components. Ultrasound faculty and the research team were up-skilled regarding ED providers’ preferences and reasons for using or not using POCUS, ways to address use barriers, and opportunities to facilitate POCUS use. Together, we reviewed intervention components that had been implemented since the last meeting, offered suggestions for improvement, outlined upcoming intervention components for the next month, and summarized the discussion. Each proposed idea was openly discussed with the entire co-creator group present, refined, and adapted based on input. Disagreements were decided upon by a majority vote. Finally, we created an evaluation and feedback plan to assess our co-design and implementation process. Updates would be shared with all ED providers at monthly faculty meetings and disseminated via email for feedback (<u>E</u>valuation).

#### 3. Data gathering process for program evaluation and feedback

Throughout our co-design process, we used modified grounded theory and iterative analysis of data collected at each co-design meeting for frequent refinement of our intervention design and implementation process. Co-creators reviewed summarized data from pre-course questionnaires, interview responses, and EHR data regarding POCUS use in the current healthcare system to promote brainstorming, reflection, and new ideas to improve the educational and clinical POCUS intervention and implementation strategy. Furthermore, ultrasound faculty met with POCUS users from other departments (e.g. Hospital Medicine, Anesthesiology/Intensive Care, and Pain Medicine) to understand how they were using POCUS clinically, identify system problems and gaps, and collaborate on establishing a hospital-wide POCUS archiving system and faculty credentialing process (<u>S</u>calability).

## III. Results

For the co-design meetings, we had good representation from all co-creator groups (3 researchers, 1 ultrasound faculty, 3 ED leadership, and 5 ED provider representatives) for a total sample size of 12 participants (Table 3). These individuals served as co-creators throughout the co-design process. Eleven participants were emergency medicine-trained (EM) physicians, 1 was an internal medicine-trained (IM) physician, and ages ranged from 31-65 years old (Table 3). The co-creator retention rate for meeting attendance and frequently contributing to sessions throughout the co-design process was 100%.

### Outcomes of the co-design sessions

Seven major topics with multiple discussion points emerged from our co-design sessions regarding the proposed one-year education and clinical POCUS system intervention and implementation plan (November 2021 to October 2022) (Table 4). We describe these outcomes by session.

**Table 4:** Topics and discussion points identified during co-design meetings with ED clinical providers (co-creators)

| Topics from the project proposal discussed in meetings | Topic discussion points identified by ED clinical staff in meetings |
| --- | --- |
| A. POCUS education (knowledge and skills) | -What are the gaps in current POCUS knowledge and use at the VA ED? |
|  | -How do we train a group with diverse baseline POCUS knowledge/skills (e.g. no prior training, beginner, skilled in some exams, advanced user)?<br>-How do we define the logistics for POCUS education sessions (number of sessions, teaching format, group sizes, frequency, etc.)?<br>-What are the facilitators for knowledge retention and what POCUS resources can we suggest?<br>-What are end-users' preferences for included content (e.g. diagnostic and procedural POCUS exams)? |
| B. Increasing uptake of clinical POCUS use | -How can POCUS use be increased at the VA ED?<br>-How can radiology US use be reduced (especially on nights/weekends when US technician is not in house)?<br>-How can patient outcomes and ED times be improved?<br>-What are the perceived barriers and facilitators to POCUS use in current practice? |
| C. POCUS documentation | -What is our current POCUS process?<br>-What is the best method to save POCUS images/clips?<br>-How should we document findings in the patient chart?<br>-How can we create a simple, standardized clinical POCUS process? |
| D. POCUS archiving | -What are the pros/cons of a systemwide vs departmental POCUS archiving system?<br>-What are current barriers to saving POCUS images?<br>-How do other departments perform POCUS (best practices)? |
| E. POCUS image review and feedback | -What are ED providers' comfort with performing/teaching POCUS?<br>-How do we create a system of checks and balances?<br>-How do we define ultrasound faculty support and responsibilities for image review/feedback (e.g. feedback methods, schedule, and frequency)? |
| F. US equipment maintenance | -Who uses the ED US machines?<br>-What is our storage, cleaning, location, and stocking process?<br>-How do we address problems with out-of-service machines?<br>-How do we share machines amongst users (availability)? |
| G. POCUS program quality improvement and feedback | -How do we evaluate our POCUS intervention and success/failures of our implementation process?<br>-How do we achieve continuous reinforcement methods through positive peer feedback?<br>-How do we maximize support from ED leadership and ED clinical POCUS champions to promote POCUS use and establish a sustainable intervention?<br>-What are the best methods for results dissemination to ED providers, ED and hospital leadership, and outside POCUS users for collaboration and future improvements? |

### 1. Initial orientation and co-design planning meeting outcomes

All co-creators met to develop a co-designed curriculum of 1) topics to be covered in educational sessions and 2) optimal clinical POCUS system processes (Table 3).

#### a) POCUS education

ED ultrasound faculty with POCUS training expertise recommended holding an introductory comprehensive in-person ultrasound training session. VA hospital medicine POCUS faculty had shared with ultrasound faculty the effectiveness of their in-person similar teaching format, which was encouraging. ED providers (end-users) were excited about hands-on training in a group environment to incorporate our ED providers’ wide spectrum of POCUS skills (from no training to beginner to advanced) and elicit active participation from many ED providers.

#### Session format

Together, co-creators brainstormed best times to offer an introductory POCUS training session with rotating hands-on stations to maximize ED provider participation and active learning, with moonlighting fellows scheduled to work in the ED that day. ED ultrasound faculty also proposed leading small-group monthly in-person scanning sessions to reinforce learning and longitudinal knowledge retention, and ED providers agreed that this repetitive time-spaced learning would be high-yield.

#### Content

Study researchers asked for content input from the individual end-users (ED providers) who would be participating in the sessions (e.g. high-yield topics, clinically useful ED exams, ultrasound-guided procedure requests, etc.) End-users requested reviewing core emergency ultrasound diagnostic and procedural exams. Finally, for POCUS resources, ED educational lead faculty suggested offering online preparatory and longitudinal learning by subscribing to a self-directed comprehensive online program with videos and questions.

#### b) POCUS clinical system and data gathered

Ultrasound faculty and ED leadership met with other VA hospital POCUS users to understand current POCUS processes. They reported that POCUS use was minimal, lacked a standardized process, and an official archiving system. Hospital Medicine users performed educational scans with individuals in small groups and shared their in-person training session outline. Anesthesiology/intensive care users were performing cardiac and lung POCUS scans but also did not have a documentation method and were reporting findings in clinical notes. No archiving or faculty credentialing system was in place in any department except in Pain Medicine solely for procedural guidance. We met with BioMedical and technical support staff to set up an ED documentation process through our EHR and had network ports built in the ED but were not successful in integrating US machine network connections with the EHR for a centralized archiving system.

Thus, given these VA hospital system difficulties, ED ultrasound faculty proposed storing clinical POCUS images on US machines and reviewing them weekly, providing quality assurance feedback via secured email. ED providers and leadership agreed this would be the best and simplest feedback method. Additional clinical process updates and information would be disseminated via faculty meeting and secured email. Per ED leadership recommendations, the finalized protocols for image documentation, archiving, and POCUS reference documents would be distributed via email and maintained on a secured VA server. Finally, on-going VA hospital wide discussions and negotiations to establish a hospital-wide POCUS archiving system would continue in parallel.

#### c) POCUS program evaluation and feedback

Co-creators jointly decided that we would obtain both individual and group feedback via monthly faculty meeting, emails, and in-person discussions between the co-creators for continuous improvement throughout the intervention and implementation process. In addition to pertinent POCUS and radiology ultrasound EHR data, study researchers would prototype and pilot pre/post-course questionnaires and interviews assessing ED providers’ POCUS skills/comfort and to obtain program feedback. Finally, ED leadership recommended using the same existing strategy for important intervention information dissemination--briefing ED providers about the upcoming POCUS course and clinical POCUS process changes via emails and at monthly ED provider meetings--to maximize acceptability and awareness.

### Co-design meeting #2

This meeting focused on developing and solidifying the educational POCUS plan. Identified topics are discussed below.

#### Topic A. POCUS educational course and knowledge retention

ED providers reported a strong desire for POCUS education.[6–8] End-users had many positive suggestions for molding the teaching sessions. They believed that tailoring educational interventions to the individual ED provider’s needs would facilitate POCUS use, with repetitive sessions over a longitudinal timeline for reinforcement. They preferred hands-on sessions, small-group over large-group, real patients with pathology for advanced users, simulation models for beginners, and quick five-minute ultrasound topics during monthly faculty meetings. All co-creators agreed that identifying and incorporating these suggestions would be key in achieving high participation rates and end-user buy-in to make our POCUS intervention an accepted and effective model.

Ultrasound faculty, lead educational faculty, and ED providers described POCUS as standard-of-care and a critical topic in resident education, citing prior ultrasound studies from the literature.[21,22] Thus, ED providers requested hands-on training time, especially for procedures such as peripheral intravenous access (US-PIV), central lines, paracentesis, and common diagnostic uses (such as cardiac, lung, fluid status, soft tissue, and musculoskeletal) in the training and educational sessions. By tailoring sessions to individuals’ needs, co-creators perceived that we would maximize POCUS uptake with sustained co-creator engagement throughout the intervention.

### Co-design meeting #3

This meeting focused on solidifying the clinical POCUS plan. Identified topics are discussed below.

#### Topic B. Increasing uptake of clinical POCUS use

Co-creators (ED providers) identified a need for a convenient and efficient clinical POCUS system with few steps involved because they perceived time constraints and system complexity as POCUS use barriers. ED providers reported bedside POCUS could improve patient care and ED flow, especially during non-business hours when the ultrasound technician is not on site.

#### Topic C. POCUS documentation

ED providers agreed with ultrasound faculty that a standardized POCUS documentation process was needed. Some providers were not familiar with how to save images on the ultrasound machines and wanted additional training on our site’s specific ED machines. All parties agreed on the need for a simple and clear documentation process that would not impede workflow or create additional burdens.

#### Topic D. POCUS archiving

Together with ultrasound faculty, end-users desired increased POCUS use and decreased radiology ultrasounds, but they did not have a good solution for image archival/documentation, and many felt uncomfortable relying on POCUS scans alone to make clinical decisions.

#### Topic E. POCUS image review and feedback

ED providers requested a formal image review process by ultrasound faculty for all clinical ED-performed POCUS scans and a quality improvement and feedback process. Ultrasound faculty introduced the concept of weekly ED POCUS image review and the importance of saving and properly labeling images for improved patient care and interpretation by reviewers, as is standard-of-care at other EDs. Ultrasound faculty also offered in-person feedback while on shift to improve both ED provider image acquisition and interpretation skills.

#### Topic F. Ultrasound equipment and maintenance

All co-creators agreed in replicating prior standardized processes to reduce cognitive load, keep systems simple, and streamline steps for POCUS use. For example, VA ED leadership and support staff would decide on ultrasound machine location by docking station outlets, space availability, and ease of access. ED nursing leadership and the ED Chief assisted with decisions on equipment stocking both with the machines and the ED stockroom given they made those decisions for ED equipment procurement, storage, and cleaning procedures.

### 3. Proposed evaluation and feedback process outcomes

#### Topic G. POCUS program quality improvement, sustainability, and feedback

ED leadership, lead educational faculty, and VA ED providers believed that information dissemination would reach the largest number of ED providers via monthly faculty meeting as a secured space for important group information, with reinforcement of material through short written email reminders. Together, the co-creator group also decided which information would be important to disseminate, and final approval was given by the VA ED chief. ED providers believed that monthly POCUS initiative updates and reminders would elicit frequent feedback and improvements based on participant input, enforcing program sustainability.

### Co-design meeting #4: Refining the multifaceted POCUS intervention through co-design and final design

Our final educational and clinical POCUS systems intervention is the collaborative product of our four co-design meetings, and the components are outlined in Supplementary Table 1. Preliminary outcome data and estimated program costs are presented in Table 5 and suggest successful program implementation, with a four-fold increase in clinical POCUS performance (72-->267 scans, p<0.001) and unchanged radiology ultrasounds (355-->361 scans, p=0.417) in the six months pre/post-intervention.

**Table 5:**
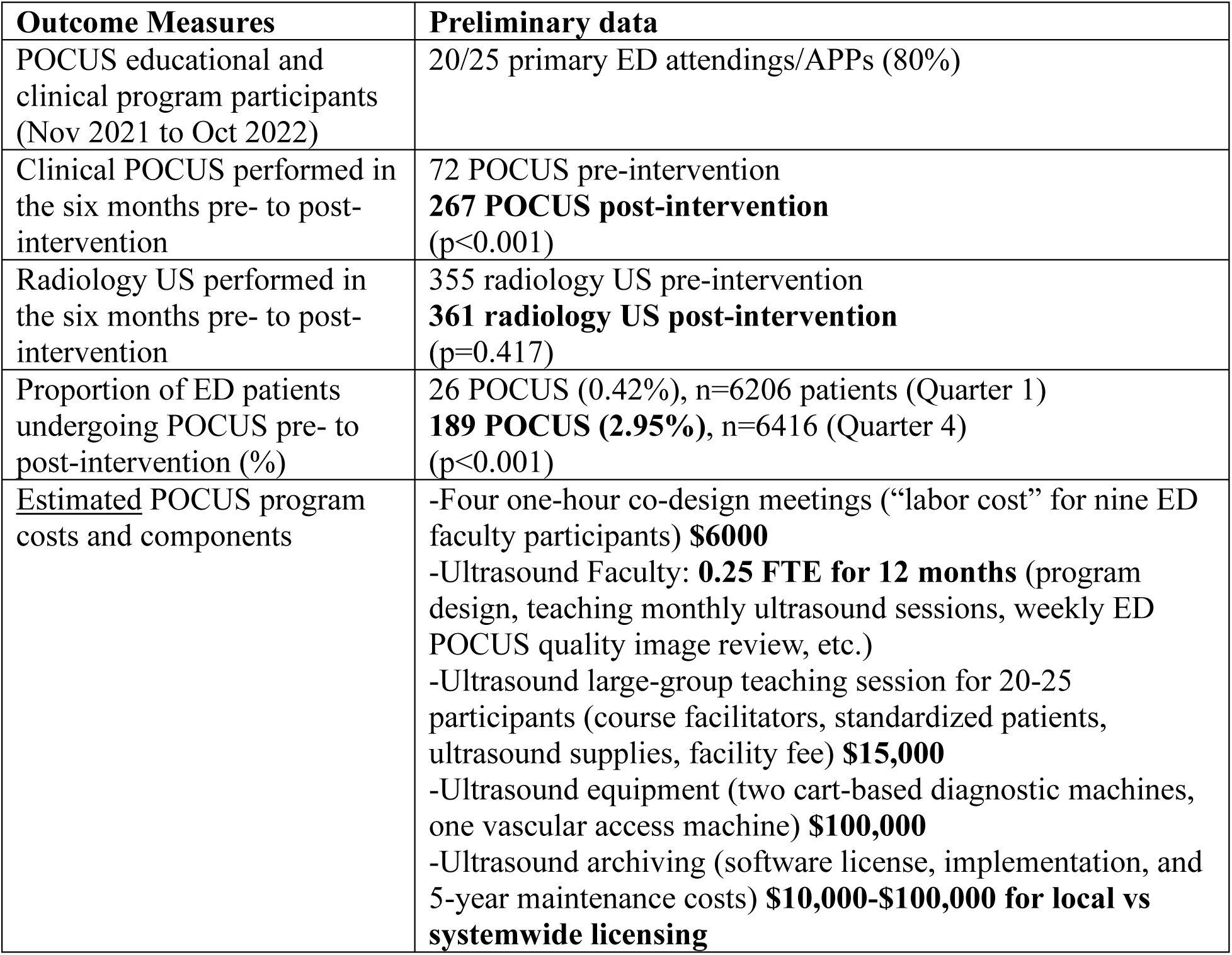
Preliminary POCUS program outcomes and estimated program costs.

## V. Discussion

Collaboration between co-creators in co-designing the intervention and implementation strategy through an iterative process allowed ED providers to help select the POCUS content taught, educational training session structure, and finalized steps required for the clinical POCUS system. Like Bird’s study that involved close collaboration between healthcare providers and family members to co-design, develop, and test a virtual care program for medically complex children and family members to better integrate healthcare at home, we structured our co-design process using PAR and EBCD methodology to streamline communication and care processes amongst team members (Figure 1).[18]

We used Leask’s PRODUCES framework to identify POCUS implementation and use problems. Preparatory planning steps with meetings between ultrasound faculty, ED leadership, other hospital POCUS users, and BioMedical technical support staff were key. By understanding the current POCUS system gaps and limitations and to underlie the foundations for our POCUS intervention, we were able to present and brainstorm POCUS system solutions in our four co-creator meetings to maximize user acceptability, uptake, and sustainability. The sequential monthly co-design meetings created progressive user feedback for adaptation of educational content and improving clinical POCUS workflow barriers and facilitators.[9]

By empowering members of the full co-design team, both VA ED leadership and ED providers collaborate as a team to achieve the best outcomes for all parties involved.[27] Rather than a one-time intervention, we created a co-designed longitudinal POCUS learning experience for ED providers to achieve sustained POCUS knowledge and utilization. Other healthcare innovation studies involving VA mental health patients and addressing problems with hospitalized veterans’ medications at transitions of care with processes similar to our study have been successful in achieving sustainable solutions.[14,28,29] Moreover, the repetitive cyclical nature of PAR and our co-designed process allows for adaptability and continuous growth and collaboration between team members throughout the intervention, which strengthens relationships and helps achieve sustainability. Our process compares with other VA studies using participatory design to improve healthcare resource access for veterans by creating an integrated health information technology system and web-based platform for veterans with mental health needs. Those studies differ from ours in their outpatient setting with patients, rather than in-hospital with healthcare providers for our study.[30–31] Other studies also support designating an ED clinical champion and note the crucial role of leadership involvement in co-design to achieve program success.[5,32] By involving co-creators throughout the design process, they feel that their views are indispensable and are empowered, yielding intervention ownership and sustained outcomes.[13,19]

To summarize, VA ED co-creators highlighted the importance of team communication, collaboration, and adaptability to overcome educational POCUS system barriers and facilitate clinical flow integration. We modeled our co-design process based on PAR and EBCD studies in the literature that defined and refined an intervention, such as Hill’s study co-designing an interdisciplinary team-based intervention for initiating palliative care in pediatric oncology, through four major co-design sessions. Their study differed from ours by creating three different interventions to better suit their three pediatric oncology teams, and they contrastingly included social workers and outside professional resources for interprofessional training.[16] Similarly, Crowther used adapted EBCD principles to develop and refine an intervention to improve guideline-adherent asthma care. The study differs by its primary and secondary care setting involvement and inclusion of both healthcare staff and patients, whereas our study focused on ED providers in a single VA ED and did not solicit patient input.[17] Like Hill’s and Crowther’s studies, VA ED co-creators can capitalize on the malleability of our intervention and implementation plan design by both anticipating potential problems and responding to difficulties throughout the process.[16–17] Finally, the iterative PAR and EBCD cycles evaluate the co-design process with continuous co-creator feedback.[9,15] The simple format of four co-design sessions with clearly defined steps and data collection methods makes this a simple and streamlined process for scalability and implementation at other sites.

### Limitations

A single standardized co-design method for designing healthcare services or products does not exist, and there are many possible frameworks and methodologies. We chose and adapted existing frameworks that best fit our setting and goals, but a different methodology may have yielded different results. Our convenience sampling method for recruitment may introduce bias since providers interested in improvement processes may not represent the entire group. To avoid bias, participation was completely voluntary, we invited ED providers of all medical specialty training backgrounds and at varying levels of POCUS use and expertise, and we ensured adequate representation from all co-creator groups at each co-design session. Furthermore, there is not a clear way of evaluating co-design methods for their outcomes. Finally, co-design can be context dependent, thus methods that work well in one context may not work at another site.[10] Nevertheless, other studies demonstrate the importance of end-user and stakeholder input in achieving an acceptable healthcare product or system process.[13,19]

### Conclusion

We describe a user-centered co-design process to maximize POCUS uptake, sustainability, and use retention in the implementation of our POCUS program and systems intervention at a single VA ED. The co-design methods described are straightforward, generalizable, and can be used as a standard framework for future POCUS program implementation and dissemination.

## Supporting information

Supp Table 1 Proposed POCUS intervention

## Data Availability

All data produced in the present study are available upon reasonable request to the authors.

