## Supplementary material for "Co-designed point-of-care ultrasound program development and implementation at a Veterans Affairs Emergency Department": Supp Table 1 Proposed POCUS intervention

**Supplementary Table 1: Final proposed POCUS intervention and implementation strategy**

| |
|---|
| *A. Final design of the POCUS educational interventions*:<br>• We will train ED providers on basic ultrasound knowledge via one 5-hour introductory half-day session, as proven by similar study methods in basic ultrasound education, followed by monthly in-person reinforcement teaching sessions.[22-26]<br>• For the introductory training course, ultrasound faculty will present an overview lecture on certain core POCUS exams, followed by hands-on training with virtual simulation devices and ultrasound scanning of healthy volunteers, divided into four one-hour rotating stations.<br>• There will be a final station with ultrasound peripheral intravenous (PIV) phantom trainers.<br>• Providers also will be given optional pre-course educational materials via a self-paid comprehensive online POCUS program subscription, which can be used independently throughout the year. |
| *B. Final design of clinical POCUS documentation and increasing use uptake*:<br>• Based on co-design meeting discussions, we jointly decided that POCUS clips and images should be labeled and saved onto the ED ultrasound machines using the patient's last name, VA patient identification number, and the performing ED provider's initials.<br>• POCUS findings will have written EHR documentation in the clinical provider's note.<br>• Also, ED ultrasound faculty will create a resource guide of required images and/or video clips for each diagnostic or procedural scan (e.g. aorta, biliary, cardiac, renal, lung, etc.)<br>• To keep ED processes standardized, the resource guide will be emailed to all providers and uploaded to a secured folder within the VA network per the VA ED Chief recommendation, as is done with other important ED protocols. |
| *C. POCUS archiving*:<br>• ED ultrasound faculty met with the ED chief, VA POCUS users in other departments (e.g. Hospital medicine, Intensive care, and Anesthesiology), our local BioMedical department faculty, VA POCUS users at our neighboring VISN site, and ultrasound vendor representatives (all stakeholders) to establish a POCUS archiving system for our VA hospital.<br>• Negotiations and approval through our local VA departments are ongoing, thus image archival is not active.<br>• Given the current lack of a standardized POCUS archiving system at our VA site and to remain consistent with ED providers' wishes for simplicity, the finalized implemented protocol is to save POCUS images on the ED ultrasound machines and document findings in the clinical note. |
| *D. POCUS image review*:<br>• ED ultrasound faculty will review all VA ED clinical POCUS scans weekly.<br>• They will provide feedback to ED providers via email if a discrepancy is noted and provide tips on improving image quality or acquisition techniques (e.g. too much depth for a biliary view, inaccurate measurement for an aorta, etc.) |
| *E. Equipment maintenance*: |

- Regarding ultrasound equipment and storage, ED providers as a group supported the decision to properly care for our machines.
- All ED ultrasound machines will be stored at the designated docking stations located in the rear of the ED, plugged in, and charged so that they are ready for use.
- Operational manuals, ultrasound gel, PIV materials, and cleaning wipes are either stored with the machines or stocked in the main ED storage room by nursing and ICT scouts.
- Machines will be cleaned between each use only with wipes approved by the machine vendor for safety.

*F. POCUS program quality improvement and feedback:*

- ED leadership will permit ED ultrasound faculty and the research team to report pertinent POCUS intervention updates to end-users at monthly ED provider meetings for continued reinforcement, reminders, or refinement, and to obtain user feedback.
- Also, with the support of the ED Chief and Deputy Director, ED ultrasound faculty will share qualitative and quantitative de-identified results at ED provider meetings, via email, and via virtual meetings to POCUS users in other VA departments.
- Lastly, ED ultrasound faculty will present results at national conferences and publish in scientific journals for dissemination to the greater ultrasound and EM community (Scalability).

*G. Plans for evaluating the new POCUS process:*

- Qualitative and quantitative findings for our POCUS intervention will be described in subsequent papers.
- Briefly, through our co-design sessions, co-creators jointly decided on a comprehensive multi-level evaluation process, including identifying the barriers and facilitators to POCUS use by our site's VA ED providers and assessing program acceptability, feasibility, and effectiveness using semi-structured interviews, questionnaires, and EHR data over a 12-month period (November 2021-October 2022).
- By following the streamlined methods used in our study with reflection at each step, the process can be replicated and implemented at other sites (Evaluation and Scalability).
